# X-ray Dark-Field Chest Imaging can Detect and Quantify Emphy-sema in COPD Patients

**DOI:** 10.1101/2021.01.15.21249798

**Authors:** Konstantin Willer, Alexander Fingerle, Wolfgang Noichl, Fabio De Marco, Manuela Frank, Theresa Urban, Rafael Schick, Alex Gustschin, Bernhard Gleich, Julia Herzen, Thomas Koehler, Andre Yaroshenko, Thomas Pralow, Gregor Zimmermann, Bernhard Renger, Andreas Sauter, Daniela Pfeiffer, Marcus Makowski, Ernst Rummeny, Philippe Grenier, Franz Pfeiffer

## Abstract

**Background:** Diseases of the respiratory system are leading global causes of chronic morbidity and mortality. While advanced medical imaging technologies of today deliver detailed diagnostic information, a low-dose, fast, and inexpensive option for early detection and/or follow-ups is still lacking. Here, we report on the first human application of a novel modality, namely X-ray dark-field chest imaging, which might fill this gap. Enabling the assessment of microstructural changes in lung parenchyma, this technique presents a more sensitive alternative to conventional chest X-rays, and yet requires only a fraction of the dose applied in computed tomography (CT).

**Methods:** For this first clinical evaluation, we have built a novel dark-field chest X-ray system, which is also capable of simultaneously acquiring a conventional thorax radiograph (7 seconds, 0·035 mSv effective dose). Representing a major medical condition, we selected chronic obstructive pulmonary disease as study subject to obtain a first impression of potential diagnostic benefits relevant to humans. For a collective of 77 patients with different disease stages, X-ray dark-field- and CT-images were acquired and visually assessed by 5 readers. In addition, pulmonary function tests were performed for every patient. The individual data sets were evaluated in a statistical work-up using correlation testing, rank-based analysis of variance, and pair-wise post-hoc comparison.

**Findings:** Compared to CT-based parameters (quantitative emphysema: ρ=–0·27, p=0·0893 and visual emphysema: ρ=–0·45, p=0·0028), the dark-field signal (ρ=0·62, p<0·0001) yields a stronger correlation with diffusion capacity in the evaluated collective. Emphysema assessment based on dark-field chest X-ray features yields consistent conclusions with findings from visual CT image interpretation and shows improved diagnostic performance in comparison to conventional clinical tests characterizing emphysema.

**Interpretation:** X-ray dark-field chest imaging allows the diagnosis of pulmonary emphysema as it provides relevant information representing the structural condition of lung parenchyma. Significant diagnostic benefits are also expected for other lung disorders.

**Funding:** European Research Council, Royal Philips, Karlsruhe Nano Micro Facility.

**Research in context:** *Evidence before this study:* With a rising number of examinations in the last decades, X-rays play an indispensable role in clinical routine. Contrast formation in medical X-ray imaging such as radiography, fluoroscopy, and computed tomography is based on attenuation, which generally benefits from large differences in atomic number and/or mass density between involved materials. If these conditions are not prevalent, or the resolution of the imaging system is not sufficient, diagnostic capabilities are limited. However, attenuation is not the only physical effect X-rays are subjected to when penetrating matter. Variations in an object’s electron density lead to refraction and coherent small-angle scattering of incident X-rays. Phase-sensitive imaging techniques can detect these wave-optical phenomena, yielding additional object information. The dark-field signal, being a function of small-angle scattering, can provide structural information on the micron scale, generally below the resolution limit of the imaging system. Due to their very stringent requirements to X-ray source coherence, these techniques were originally limited to large-scale synchrotron facilities. The proposal of a three-grating interferometer in 2006, however, enabled the use of low-brilliance sources for X-ray phase-contrast imaging and thereby paved the way into the clinics. Such an apparatus elegantly allows the simultaneous acquisition of the conventional attenuation, differential phase-contrast, and novel dark-field signals. In a compact table-top system suitable for investigating murine disease models, numerous studies on pulmonary disorders such as chronic obstructive pulmonary disease (COPD), pulmonary fibrosis, pneumothorax, ventilator-associated lung injury, lung cancer, and pneumonia have been conducted and demonstrated a broad diagnostic value of the dark-field modality in particular. Adapting the system to enable imaging of the human body is a technical challenge due to limitations of the micrometer-fine, high aspect ratio grating structures in terms of fabricable size and performance at clinically relevant X-ray energies. The first evidences that these limitations are manageable were delivered in 2017 and 2018 by in-vivo porcine and human cadaver studies with an experimental prototype system.

*Added value of this study:* With this work we present the first X-ray dark-field chest images of human subjects in-vivo and demonstrate the method’s feasibility in a clinical surrounding. To enable this study, we have conceived, constructed, and commissioned a custom-built first demonstrator system suitable for patient use. This includes satisfying clinical demands regarding safety, usability, acquisition time, radiation dose, field of view, and image quality. This study marks the transition from investigating artificially induced disease models to evaluating the modality’s actual diagnostic performance in patients.

*Implications of all available evidence:* Our findings indicate that X-ray dark-field radiography provides image-type information of the lungs’ underlying microstructure in humans. In view of the strong link between alveolar structure and the functional condition of the lung, this capability is highly relevant for respiratory medicine and might help to establish a better understanding of pulmonary disorders. With regard to early detection of COPD, which is generally accompanied by structural impairments of the lung, this novel technique might support resolving the prevalent under-diagnosis reported in literature. With an effective dose significantly lower (about a factor of hundred) compared to thorax computed tomography, dark-field radiography could be used as broadly deployed screening tool.

## Introduction

Although diagnostic imaging systems have seen tremendous technological advances over time, assessment of pulmonary microstructural changes remains challenging in clinical routine. Here, we report on the first human in-vivo application of a novel radiographic modality, namely X-ray dark-field chest imaging. Introduced on the lab bench in 2008,^1^ this technique delivers complementary imaging information on the lung’s microstructure beyond conventional chest X-rays.^2–4^ In contrast to common X-ray imaging, which measures the attenuation of X-rays, dark-field contrast is related to coherent small-angle scattering of X-rays at microscopic structures within the specimen.^1,5^

The lung is composed of very fine interfaces between air and soft tissue in order to maximize the surface to volume ratio, which is essential for an efficient gas exchange. Repeated refraction and scattering of incident X-rays at these interfaces induces a high dark-field signal, which constitutes the great potential of the technique with respect to pulmonary imaging^2–4^. Pulmonary disorders such as emphysema, fibrosis, lung cancer, or pneumonia have direct impact on these structural properties by degrading, condensing, displacing, or infiltrating lung tissue, each leading to an alteration of the dark-field signal. With regard to these and further conditions, its diagnostic superiority over conventional chest radiography has been demonstrated in various small-animal studies.^6–12^

The aim of this work is to demonstrate the first successful application of dark-field chest radiography in living humans and, at the same time, to highlight the technique’s future potential for diagnostic imaging of human lungs. We selected pulmonary emphysema in patients with chronic obstructive pulmonary disease (COPD) as the subject of the first experimental trial on humans, not least because of the strong supporting evidence of the technique’s benefits found during several small-animal dark-field imaging studies on emphysema detection and staging.^2,3,6,7^ COPD represents a major medical condition claiming millions of lives every year worldwide.^13^ It is a chronic inflammatory process of the airways often involving destruction of alveolar and vascular structure, characterized by irreversible airflow limitations. Representing one component of COPD, pulmonary emphysema is characterized by permanent dilation of air spaces and destruction of their walls distal to terminal bronchioles. Computed tomography (CT) is a validated imaging technique to visually and quantitatively assess the presence, extent, and pattern of emphysema in-vivo. In addition, for subjects with established COPD, chest CT provides data on the risk of lung function decline, respiratory exacerbations, and death, independently and incrementally to routinely used studies, such as pulmonary function tests and symptom measures.^14^ Chest CT, however, is currently not considered as standard of care in the diagnosis and management of mild to moderate COPD.^15^ In particular, the suitability of CT is restricted by high radiation exposure of about 7 mSv^16^ for a regular chest examination. Even if low-dose techniques are applied, the radiation exposure is still in the range of 2 mSv.^17^

A chest X-ray is usually the first diagnostic tool used in the evaluation of the patient’s lungs, because it is fast, accessible, and inexpensive. The applied radiation dose is comparably low with an average effective dose of about 0·02 mSv for a single posterior-anterior image.^16^ However, it is well documented that plain chest radiography has low sensitivity for detecting emphysema.^18^

Because of its demonstrated high sensitivity to microstructural parameters, the question arises whether the novel dark-field technique may offer an alternative low-dose imaging approach that allows an improved medical assessment of the lungs. In order to address this question, we have conceived, constructed, and commissioned a custom-built first demonstrator system suitable and certified for human use within a university hospital (Klinikum rechts der Isar, Technical University of Munich). In this manuscript, we report initial results regarding the diagnosis of COPD based on a collective of 77 patients and compare X-ray dark-field imaging with pulmonary function tests, symptom measures, and medically indicated chest CT.

## Methods

### X-ray dark-field demonstrator system

In order to record the first clinical dark-field chest X-ray images in humans, a clinical demonstrator system capable of acquiring both dark-field and attenuation X-ray images of an entire human thorax in about 7 seconds scan time was conceived, constructed, and integrated in the Department of Radiology. The present implementation of the system represents the final fusion of several important technological developments, which have been pursued in multiple preparatory research projects leading up to this work.^1,4,19–25^ Figure 1A shows a schematic of the system arrangement. The system’s design is based on standard medical components, such as X-ray source, collimator, and flat-panel detector, which are combined with a three-grating Talbot-Lau X-ray interferometer.^1,19^ The acquisition procedure is realized in scanning geometry, and uses a modified fringe-scanning method.^26,27^ Via phase modulation of the X-ray beam, the grating G_1_generates a high-frequency intensity pattern with a periodicity of a few microns at the position of the grating G_2_ that consists of highly absorbing gold lines. By utilizing a similarly high spatial frequency in the G_2_ structure, this pattern is down-converted to low-frequency moiré fringes with a period of a few millimeters, which can be recorded conveniently by conventional medical imaging detectors.

**Figure 1:**
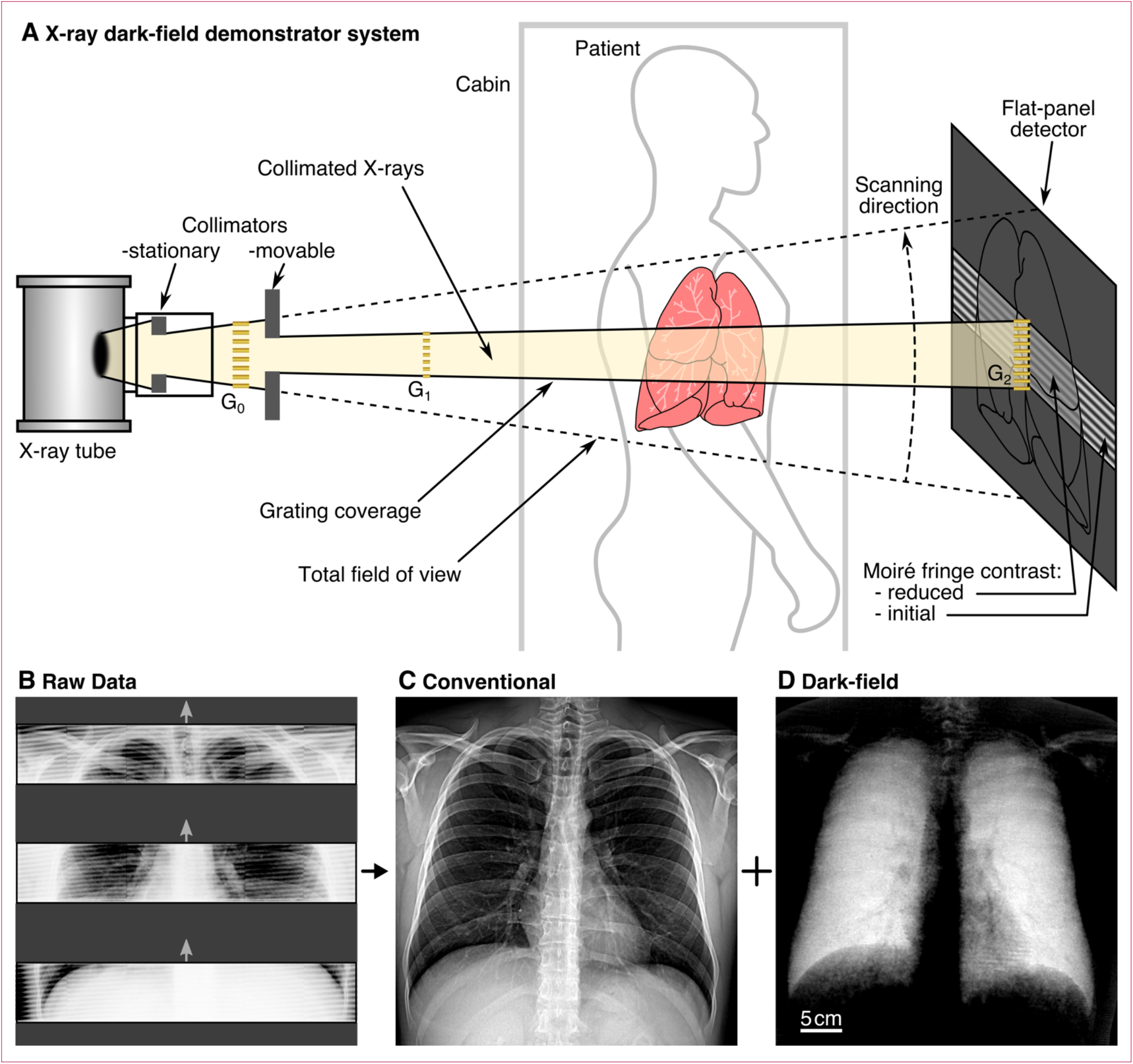
Schematic of the X-ray dark-field demonstrator system (A), exemplary raw detector imaging data (B), as well as retrieved conventional (C) and dark-field (D) chest X-rays from first in-human application. The moiré fringe pattern, which is generated by the interplay between the phase grating G_1_ and the analyzer grating G_2_, is scanned across the patient’s thorax. Subsequent processing of the image stack analyzes the changes in the moiré fringe pattern and retrieves conventional and dark-field radiographs based on attenuation and small-angle scattering of X-rays within the patient’s thorax. Behind regions with a high fluctuation of the refractive index (e.g. alveolar microstructure), the contrast of the moiré pattern is reduced as a result of multiple small-angle scattering. The grating G_0_ allows the use of standard X-ray source technology with a large focal spot. Panel B depicts three (out of 195) exemplary raw-data frames recorded during one scan of a healthy patient (male patient, 33 years, BMI 30·2, without lung disorders). The moiré pattern is clearly visible, superimposed onto the patient’s thorax, which is subsequently used to retrieve a conventional attenuation chest radiograph (C) and the novel dark-field chest radiograph (D). Intact alveolar structure with a high density of air-tissue interfaces induces strong small-angle scattering, resulting in a pronounced dark-field signal within the lung region. The magnitude of the signal correlates with the number of interfaces^7^ and thus provides structural information on alveoli that are typically smaller than the resolution limit of the imaging system. The scale bar in D also applies for B and C. Key system specifications: Field of view: 37×37 cm^2^ (patient plane); Effective radiation dose: 0·035 mSv (male, 73 kg); Acquisition time: 7 seconds.

Finally, the grating G_0_ is necessary to allow the use of high-power medical X-ray sources, which typically have rather large focal spot sizes (i.e. several hundred microns). The installation of G_0_ close to the source ensures that every individual beamlet coming from a gap in G_0_ has sufficient transverse coherence of X-rays, necessary for the formation of the high-frequency intensity pattern.^19^

If an object with a large number of inherent interfaces on the micron scale (e.g. lung tissue) is placed within this arrangement, the contrast (visibility) of the moiré pattern decreases as a result of multiple refractions on these interfaces.^2–4^ This allows assessing characteristics of structures significantly smaller than the resolution limit of the imaging system.^1^ The magnitude of the dark-field signal is encoded in the reduced visibility *V* of the moiré pattern, whereas the magnitude of the attenuation signal is given by the pattern’s decreased mean intensity *I*. Both quantities are normalized to air scan values (*I*_0_, *V*_0_) to account for inhomogeneities ranging over the field of view. As intensity and visibility decline exponentially with object thickness,^5^ both signals are here given as the negative natural logarithm of the respective normalized quantity, i.e.: – ln(*I*/*I*_0_) and – ln(*V*/*V*_0_). Both modalities are jointly calculated from the data acquired within a single scan, yielding two perfectly registered images. To cover the region of an entire human thorax, the interferometer with an oblong grating area of about 42×6·5 cm^2^ is moved across the patient, resulting in a field of view of 37×37 cm^2^ with respect to the patient plane. Simultaneously, a series of consecutive images is recorded by the flat-panel detector. Figure 1B shows three exemplary raw data frames from the examination of a patient without pulmonary disorders. An entire sequence of the scan is made available in the supplementary material online (video 1).

The data processing essentially relies on the approach described in ref.^27^, extended by additional correction and calibration procedures to account for signal corruption induced by Compton scattering and beam hardening. The moiré pattern in each pixel of the raw images is mapped onto a sinusoidal intensity model, which allows the extraction of the pattern’s mean value (figure 1C: conventional attenuation) and visibility (figure 1D: dark-field).

The lungs appear radiolucent in the conventional image, primarily related to their high air content. In the dark-field image by contrast, a distinct and homogeneous signal from the lung is obtained. Intact alveolar structure with numerous air-tissue interfaces induces strong small angle-scattering, resulting in a highlighted depiction of the lungs that allows for an unimpeded assessment.

In order to minimize radiation exposure, our system was designed with specific focus on dose efficiency. An inverse interferometer arrangement^28^ was selected to avoid the G_1_ grating acting as an additional absorber downstream of the patient position. Secondly, a combination of a stationary (MTR 302, Ralco, Italy) and movable (2 mm Tungsten) collimator was used to avoid exposure in regions that are not covered by the gratings and hence do not contribute to the image. After moving the interferometer out of the beam path, the stationary collimator can be adjusted according to the entire region to be imaged. This is achieved with motorized collimator blades that are illuminated for visual position feedback. The movable collimator is attached to the interferometer and restricts the beam to the grating area. An additional measure for minimizing patient exposure is synchronizing detector readout and exposure to avoid dose deposition during insensitive time intervals. Further, intrinsic beam filtration (2·5 mm Aluminum) removes low-energy photons that would not penetrate the patient and thus would not contribute to image formation.

Downstream of the two collimation layers covering the entire radiation area, an ionization air-chamber (Diamentor CI, PTW, Germany) is mounted to log the applied radiation dose of every scan. Via a measurement of the dose area product, the effective patient dose applied during one scan is estimated. The respective conversion coefficient (1·5 µSv/µGy.m^2^) was determined utilizing an anthropomorphic thorax phantom (ATOM 701, CIRS, USA) with embedded thermoluminescent dosimeters. The phantom simulates X-ray attenuation properties of a male reference person with a body size and weight of 173 cm and 73 kg. With respect to such a reference person undergoing one scan in posterior-anterior orientation, the received effective dose was determined to be 0·035 mSv. To ensure a constant dose in the image receptor plane (∼3·5 µGy, behind the lungs), the tube current is modified according to the patient’s size and weight. This ensures a constant image quality over the patient collective, but also implies that patients with a higher BMI receive a higher radiation dose. The acceleration voltage was set to 70 kV_p_, which is a reasonable compromise between interferometer performance and X-ray transmission.^24^

### Pulmonary function test and symptom measure

All participants underwent lung function testing according to the European Respiratory Society recommendations.^29,30^ These tests consist of a combination of spirometry with body plethysmography (MasterScreen Body, Jaeger, Germany). For 42 patients, the individual diffusion capacity of the lung for carbon monoxide uptake during one single breath (DLCO SB) was gathered additionally (MS-PFT, Jaeger, Germany).

Clinical symptoms were evaluated using a COPD questionnaire (CAT™, GlaxoSmithKline, United Kingdom).

### Computed tomography

Imaging was performed on a CT scanner (IQon Spectral CT, Royal Philips, The Netherlands) during clinical routine using a standard clinical chest protocol. The tube voltage was set to 120 kV_p_ and the system’s automatic angular tube current modulation was utilized. After a volumetric acquisition over the entire chest at full suspended inspiration, 0.9 mm-thick axial slices were reconstructed with high resolution and soft tissue algorithms (iDose, Royal Philips, The Netherlands) for visual and quantitative assessment, respectively. The vendor-specific noise suppression level 6 (iDose 6) was applied. Iodine contrast material was present for all patients due to recruitment among cancer treatment follow-ups. Quantitative analysis of emphysema was performed by thresholding lung density at –950 HU using a commercially available software (IntelliSpace, Royal Philips, The Netherlands) in order to obtain the CT emphysema index (ratio between emphysema- and lung volume). According to the Fleischner Society, CT emphysema index values below 6% were not considered to represent significant emphysema.^31^

### Visual analysis

Visual emphysema assessment based on CT images was performed by three trained radiologists. As a scale, the Fleischner Society classification scheme^31^ was applied, grading emphysema severity by the groups *absent, trace, mild, moderate, confluent*, and *advanced destructive*.

On the basis of the dark-field chest images, five readers (all radiologists with knowledge in dark-field contrast formation) graded emphysema severity for each patient on a five-point Likert-scale with the levels *no evidence of emphysema* (0), *beginning-* (1), *mild-* (2), *moderate-* (3), and *severe emphysema* (4). Diagnostic confidence for this rating was also collected (levels: 1–4). In addition, dark-field signal intensity and homogeneity was rated within upper (u), middle (m), and lower (l) subregions of the left (L) and right (R) lung, respectively. The signal intensity was graded on a seven-point Likert-scale ranging from 0 (no signal) to 6 (maximum signal). Signal homogeneity was rated applying a four-point Likert-scale with the levels *very inhomogeneous* (1), *moderately inhomogeneous* (2), *mildly inhomogeneous*/*mostly homogeneous* (3), and *homogeneous* (4). Additionally, in the case of a homogeneity rating other than *homogeneous*, areal signal texture was graded by assigning the levels *no texture* (0), *subtle stains* (1, <5 mm), *medium stains* (2, 5-10 mm), and *large stains* (3, >10 mm).

Visual assessment of CT and dark-field images was temporally decoupled and patient order was randomized. Revision and adaptation of previous grading decisions were allowed during readings.

### Statistical analysis

The Likert scales used for feature grading in the dark-field image readings were considered as interval-scaled, in order to obtain refinement between the grades by calculating the mean over all readers.

To assess the consistency of visual signal grading, the rated levels were compared to the mean dark-field signal measured over all image pixels included in the respective subregions. For this purpose, a mask was generated by manually selecting the outline of the lungs and subsequently calculating the average signal value of all included pixels. Lung regions superimposed by the heart and the diaphragm were omitted in order to exclude size-specific contributions from these parts. The mask was split into the subregions of the reader study (u, m, l) for each lung. Exemplarily, figure S3 in the supplementary material shows the respective mask for the patient depicted in figure 1B-D. Rank correlation coefficients between rated and measured signal were calculated for comparison.

In order to investigate how the functional condition of the lung is represented by CT and dark-field, correlation tests with diffusion capacity (DLCO SB) from pulmonary function testing were conducted. As test parameters, quantitative emphysema index and visual emphysema grade (reader median) both derived from CT as well as dark-field signal rating were utilized. The individual data sets were tested for normal distribution using ShapiroWilk test based on a rejection for p<0·05. As most data sets do not follow Gaussian distributions, rank correlation coefficients were used for the analysis. P-values less than 0·05 were considered to indicate significant correlations. In order to obtain a single-value measure that describes the dark-field signal strength for each patient’s lungs, the mean rated signal level over all readers and subregions (u-L, m-L, l-L, u-R, m-R, l-R) was calculated. For the assessment of the dark-field chest images’ clinical performance, the patient collective was grouped by Fleischner grade (reader median), serving as diagnostic reference for pulmonary emphysema. The groups *confluent* and *advanced destructive* were combined in the evaluations to achieve a sufficient sample size. In order to compare diagnostic accuracy, descriptive statistics within these groups were calculated for conventional clinical measures and for image features derived from the dark-field readings. In addition, parameter variation over all groups and pair-wise variation between adjacent groups were evaluated using Kruskall-Wallis (critical value at H=9·49 for a confidence level of 95%) and Mann-Whitney-U (difference was considered significant if p<0·05) tests, respectively. For the lung function parameters *forced expiratory volume in 1 second* (FEV1) and r*esidual volume*, the percentage of the expected value for a patient without pulmonary disorders having same age, sex, and height was used. The parameter for dark-field-based emphysema severity was obtained by calculating the average rating from all five readers. For the signal homogeneity and texture level, the mean over all six subregions and all readers was calculated. Open-source software frameworks (SciPy, Pandas, Python) were used for the calculations.

### Patients

The study was approved by the institutional ethics review board and the German Federal Office for Radiation Protection (Bundesamt für Strahlenschutz, BfS) and the permission from authorities to operate the demonstrator system for first human applications was obtained. A collective of 77 patients were included prospectively in the study. All participants gave written informed consent and were approached after undergoing medically indicated chest CT.

To investigate the technique’s performance in particular for early disease stages, the focus was placed on patients without pulmonary disorders as well as those with initial indications of emphysema revealed in CT, but still absent obstruction according to pulmonary function testing (FEV1/FVC>0.7). However, moderate to severe stages were also included to some extent in order to obtain an impression of the (full) signal range of dark-field signal across the entire spectrum of emphysema severity. Table 1 provides an overview of the study population.

**Table 1:** Study population (*N*=77, 30 female, 47 male). Global Initiative for Chronic Obstructive Lung Disease (GOLD) stage assignment is according to ref.^32^.

| Parameter | mean | min. | max. |  |  |
| --- | --- | --- | --- | --- | --- |
| Age (female/male) [y] | 64.9<br>(63.3/65.9) | 30<br>(42/30) | 91<br>(80/91) |  |  |
| Body mass index [kg/m<sup>2</sup>] | 26.0 | 16.4 | 38.9 |  |  |
| **GOLD stage:** | **no obstruction** | **I** | **II** | **III** | **IV** |
| No. subjects | 64 | 2 | 6 | 3 | 2 |

## Results

Figure 2 illustrates the types of data generated during the study, using an exemplary male patient (58 years, BMI 27·6, 35 pack-years of smoking). The conventional attenuation image (figure 2A) shows no conspicuous features suggesting structural impairments of the lung. Most parts of the lung yield dark-field values (figure 2B) comparable to the ones of the healthy lung in figure 1D. However, in the apical area of the right and left lung, spreading over approximately 6–7 cm in caudal direction, the signal is strongly decreased. Additionally, in the middle right zone, a moderate reduction is recognizable reaching across the prominent interface between upper and middle lobe. These observations are consistent with this case’s mean reader ratings of the dark-field signal intensity (u-R: 1·8, m-R 2·8, l-R: 4·6, u-L: 2·0, m-L: 5·0, l-L: 4·8), signal homogeneity (u-R: 1·4, m-R 1·4, l-R: 2·8, u-L: 1·4, m-L: 2·8, l-L: 2·8), and areal signal texture (u-R: 2·8, m-R 2·8, l-R: 1·6, u-L: 2·8, m-L: 1·2, l-L: 1·2). Based on the dark-field image, emphysema severity was graded between moderate and severe (3·6) on average. The air-flow curve (figure 2C) shows a characteristic concavity in the descending limb indicating obstructed airways.^33^ Except for FEV1, parameters from pulmonary function test (figure 2C) are deviating moderately from predicted values after administration of a bronchodilator. The FEV1/FVC ratio lies just at the abnormality threshold (FEV1/FVC=0·70, prior to bronchodilation) applied in the GOLD classification scheme.^32^ The still decreased FEV1 remaining post bronchodilation indicates irreversible air-flow impairment. Also, the decreased DLCO SB indicates reduced alveolar surface. In figure 2D, coronal and sagittal CT slices of the thorax are depicted. A fraction of 11·5% of all voxels assigned to the lung are classified as emphysematous, yielding density values lower than –950 HU. Notably, affected regions (marked in red) coincide with those of decreased dark-field signal. In the right lung for instance, highlighted voxels align along the inclined lobe interface, matching the region with the moderately decreased dark-field signal. In contrast, regions featuring higher density values coincide with regions of stronger small-angle scattering, indicating an intact alveolar structure in both modalities, respectively. Visual CT assessment based on the Fleischner scheme yielded *confluent* emphysema for this patient’s lungs.

**Figure 2:**
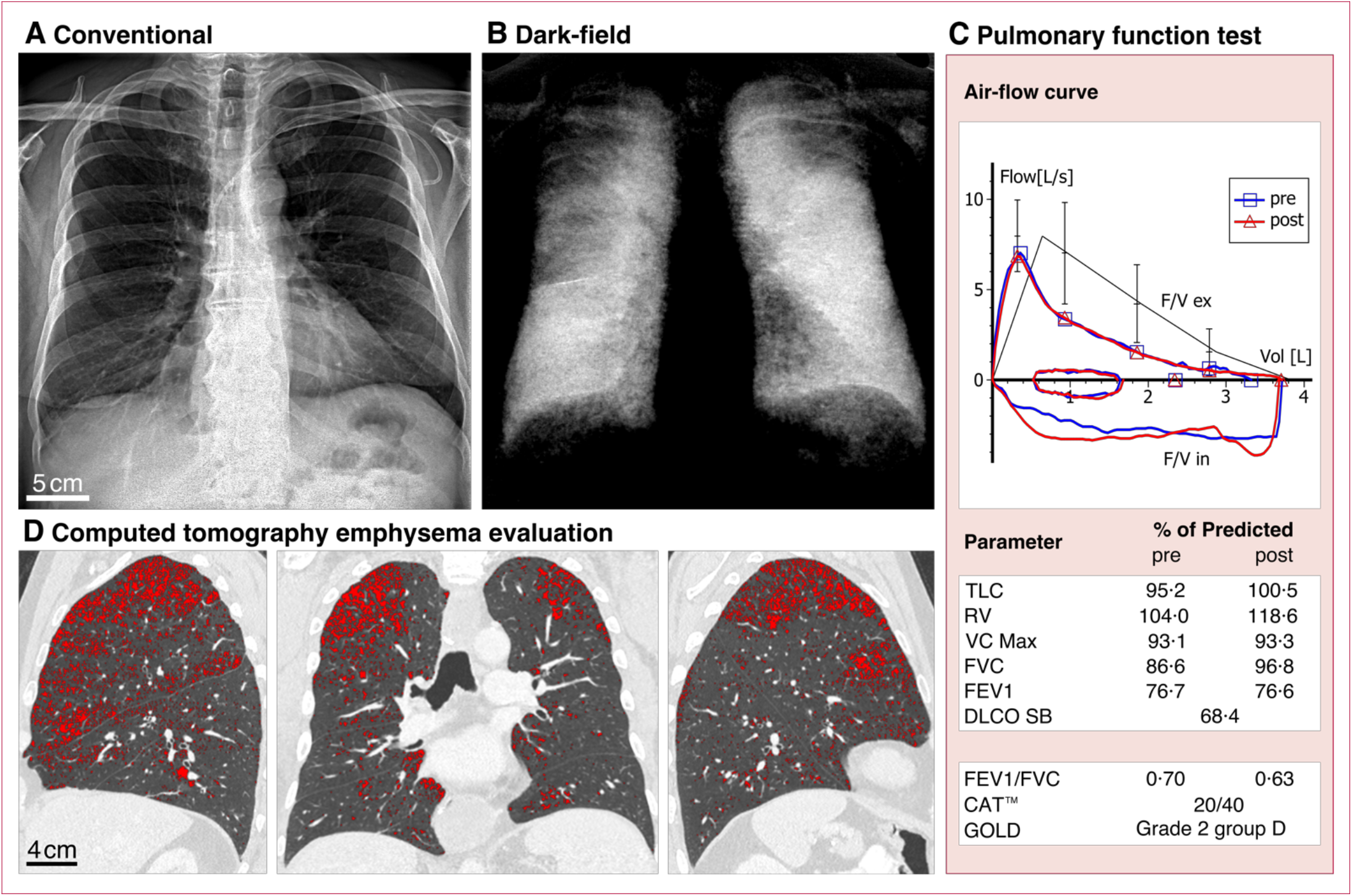
Results from imaging (conventional and dark-field chest X-rays, CT) and pulmonary function testing for an exemplary patient. (male, 58 years, BMI 27·6, 35 pack-years). Conventional (A) and dark-field (B) chest X-rays of a patient with localized emphysema, recorded with our demonstrator system. In the dark-field image (B), affected regions appear dark, indicating reduced small-angle scattering due to a decreased number of air-tissue interfaces. In contrast, the attenuation image (A) is very limited in visualizing the circumscribed emphysematous changes. Pulmonary function test (C) yields moderate obstruction still remaining post bronchodilation (Abbrev.: TLC: total lung capacity, RV: residual volume, VC Max: maximal vital capacity, FVC: forced vital capacity, FEV1: forced expiratory volume in 1 second, DLCO SB: diffusion capacity of the lung for carbon monoxide in one single breath, CAT™: COPD assessment test). Each lung function parameter is normalized with respect to the expected value of a patient without pulmonary disorders having same age, sex, and height. CT densitometry yields an emphysema index of 11·5% implying this fraction of the lung volume has density values below –950 HU. The corresponding voxels in the CT slices (D, lung parenchymal window settings applied) are marked in red. Dark-field and CT images yield consistent information with regard to parenchymal condition.

In figure 3, dark-field images (A–L) of 12 patients of our study with gradually decreasing signal intensity in the pulmonary region are lined up with the corresponding attenuation images (M– X). Table 2 lists the associated body mass index (BMI), FEV1/FVC ratio, CT emphysema index, as well as the average dark-field and attenuation values extracted from the lung region.

**Table 2:**
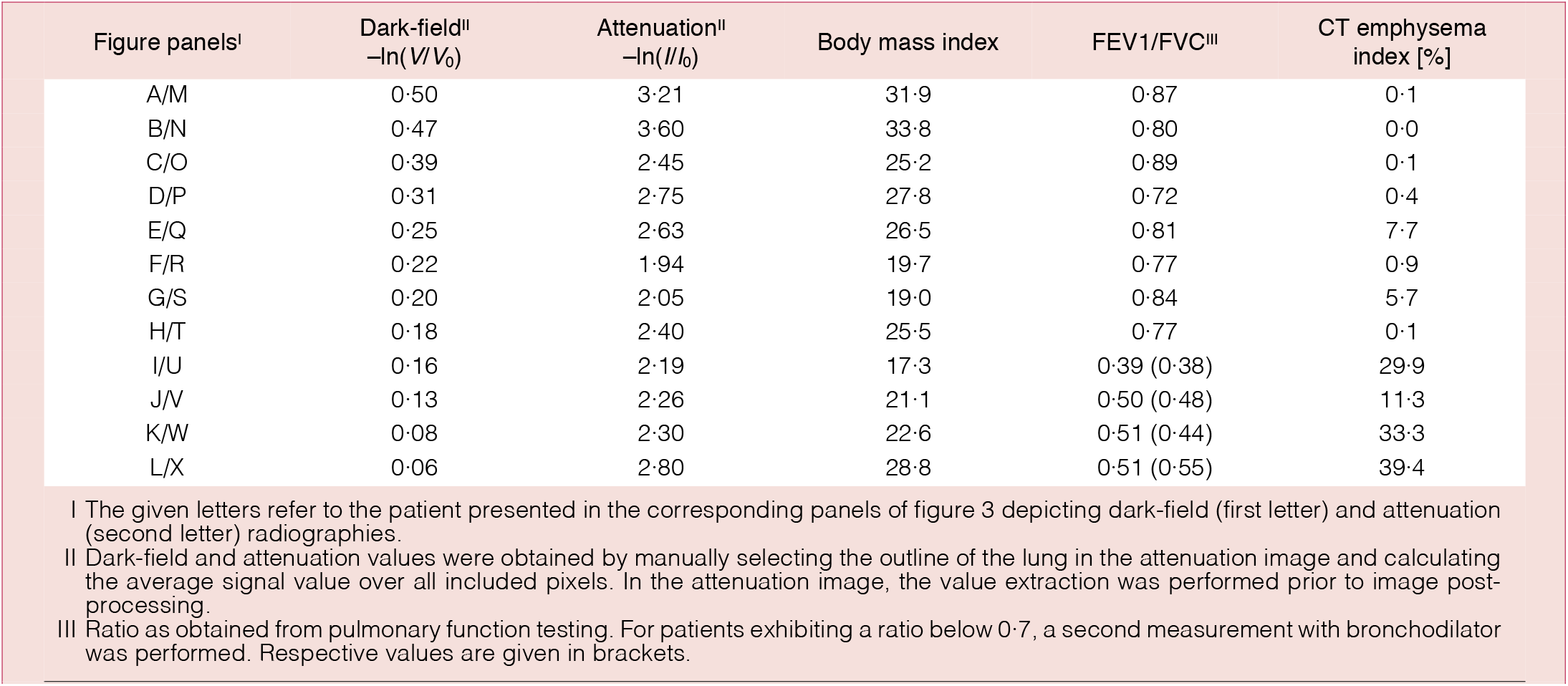
Dark-field, attenuation, spirometry, and CT findings for patients with increasing severity of COPD. In contrast to the attenuation signal, which is rather related to the BMI of the patient, the dark-field signal declines gradually with decreasing pulmonary function and increasing emphysema index.

**Figure 3:**
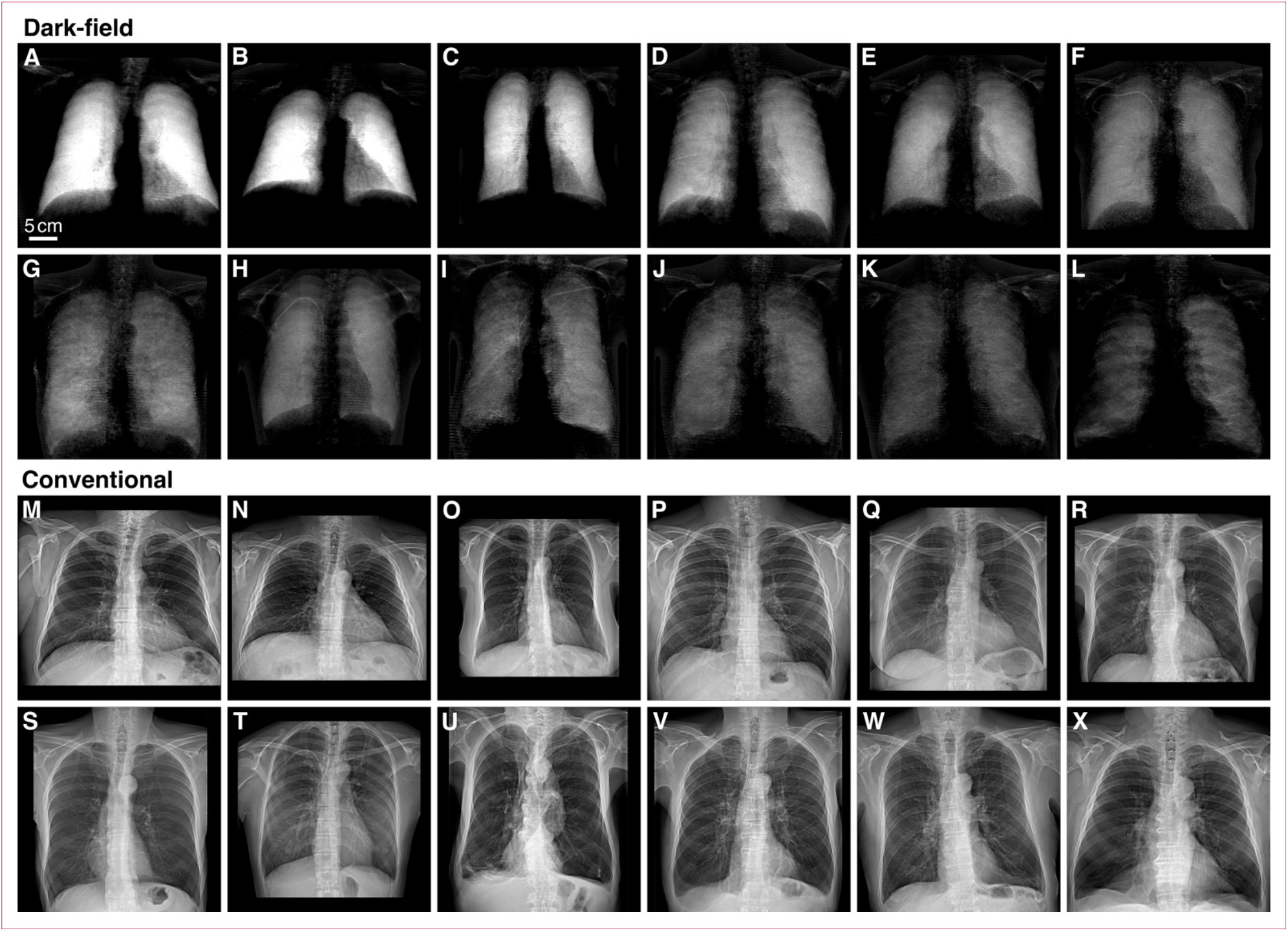
Dark-field and conventional chest X-rays for patients with increasing severity of COPD. In the dark-field images (A-L), a gradual decrease in signal strength, ranging from no pulmonary disorder to severe emphysema, can be identified. In addition, local signal variations can be recognized. With a decline in the number of alveolar interfaces, less small-angle scattering is induced, resulting in decreasing dark-field signal values. No significant variation in contrast of the lung relative to surrounding features is recognizable in the respective attenuation images (M-X). The ribcage, superimposing the lungs in the attenuation image, is barely present in the dark-field images. Patient sorting: The dark-field/attenuation pairs A/M, B/N, C/O, D/P, E/Q, F/R, G/S, H/T, I/U, J/V, K/W, and L/X correspond to the same patient. Within the modalities, identical window and level settings are applied. The scale bar in (A) applies for all panels.

In this group, a decreasing dark-field signal is accompanied by decreasing pulmonary function and increasing CT emphysema index, which are characteristic indications for a reduction of the alveolar surface. In the attenuation images, the observed variation in dark-field signal is not accompanied by notable contrast variations of the lung with respect to the surrounding area. The average signal within the lungs is rather related to the collective attenuation by all materials overlaying the lungs, in particular muscle, fatty tissue, and breast tissue. This is reflected by a very strong correlation between mean attenuation signal and BMI (r=0·84, p<0·0001, *N*=77). Apart from effects such as Compton scattering and beam-hardening, which are correctable to the largest extent, the dark-field signal is not affected by microscopically uniform and thus non-scattering material superimposing the lungs. While the dark-field signal exhibits a gradual decrease, inconspicuous findings for the FEV1/FVC ratio (8 patients, FEV1/FVC > 0·7)^32^ and emphysema index (7 patients, CT emphysema index < 6%)^31^ were obtained among these 12 examples.

Indicating consistent reader grading, strong positive correlations were found between measured and rated dark-field signal intensity for all subregions (u-R: 0·76, m-R: 0·86, l-R: 0·87, u-L: 0·75, m-L: 0·84, l-L: 0·85, all p<0·0001, *N*=77).

Visual emphysema grading based on the CT images yielded 35 patients with *absent-*, 19 with *trace-*, 9 with *mild-*, 7 with *moderate-*, 6 with *confluent-*, and 1 with *advanced destructive* emphysema.

Figure 4A shows scatter plots of DLCO SB versus CT emphysema index, visual emphysema grading, and rated dark-field signal intensity for all patients who underwent an examination of the diffusion capacity (*N*=42). The dark-field signal yields a stronger correlation (ρ=0·62, p<0·0001) compared to the two CT-based parameters (ρ=–0·27, p=0·0893 –not significant and ρ=–0·45, p=0·0028). Considering only the patients in the Fleischner groups *absent-, trace-*, and *mild emphysema* (*N*=35), for these correlation pairs we find no significant correlation for the CT parameters (ρ=–0·07, p=0·7000 and ρ=–0·31 p=0·0681) and a moderate correlation for the dark-field signal (ρ=0·46, p=0·0056). In addition, we find moderate correlations for CT emphysema index, visual emphysema grading, and rated dark-field signal with FEV1/FVC (ρ=–0·52, ρ=–0·53 and ρ=0·49; all p<0·0001, *N*=77).

**Figure 4:**
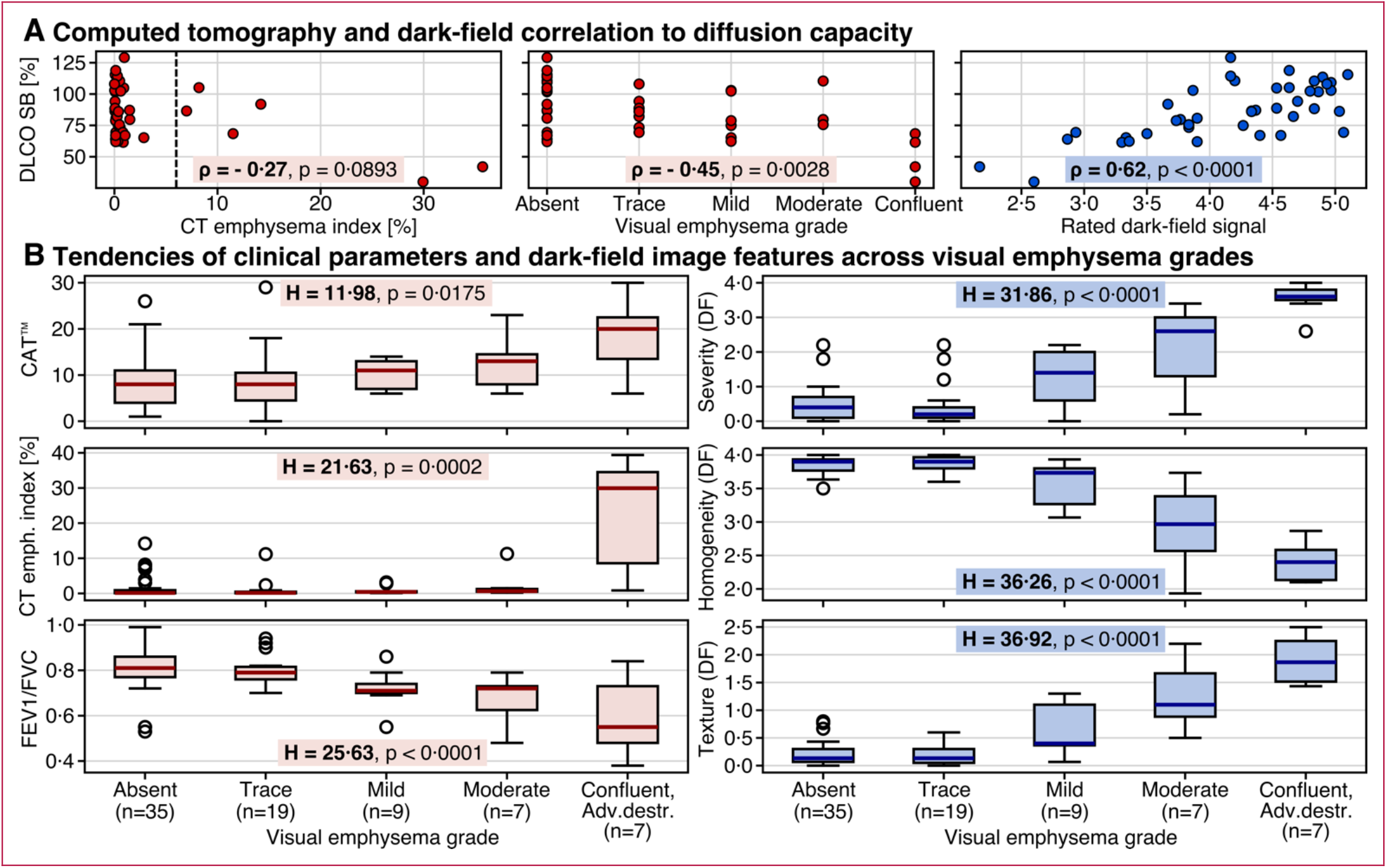
Main results from study evaluation. The scatter plots (A) illustrate the respective correlations of the CT emphysema index, CT-based visual emphysema grade and rated dark-field signal intensity with the diffusion capacity of carbon monoxide (DLCO SB). For the latter, values are given with respect to the expected value of a patient without pulmonary disorders having same age, sex, and height. Visual emphysema grades correspond to the median rating of the three readers. Values for rated dark-field signal are means over all six subregions and all five readers. The strong link between diffusion capacity and structural lung condition is reflected in the good correlation with the dark-field signal: Both DLCO SB and dark-field signal describe the condition of the respiratory surface with the ability to perform gas-exchange and small-angle scattering at microscopic interfaces, respectively. In (B), box plots illustrate the tendencies found for the CAT™ score, CT emphysema index, FEV1/FVC ratio, dark-field-based emphysema rating and dark-field image features across the Fleischner emphysema grades of the evaluated collective. If FEV1/FVC<0·7, the measurement post bronchodilation was utilized for the FEV1/FVC ratio in the evaluation. The parameter for dark-field-based emphysema severity corresponds to the mean over all five reader ratings. To obtain parameters for homogeneity and texture level, the average over all six subregions and all readers was calculated. The Fleischner grade is assigned based on the size and distribution of regional density variations induced by parenchymal impairments. The associated alteration of alveolar microstructure also induces local signal variations in the dark-field image due to reduced small-angle scattering in affected regions. Consequently, a decreasing signal homogeneity and increasing texture is observable in the dark-field images with increasing visual emphysema grade.

Table 3 gives an overview about the study results. Rank-based analysis of variance indicates significant value variation across the Fleischner grades for all evaluated parameters, conventionalclinical and dark-field based emphysema measures. Post hoc pair-wise comparison of adjacent groups (*absent–trace, trace– mild, mild–moderate, moderate–confluent/adv. destructive*), however, yields improved discriminability for the visual features in the dark-field images compared to the conventional measures for emphysema diagnosis: Of in total four, none of adjacent group pairs showed significant differences in the CAT™ score. For the parameters from pulmonary function testing, we found one group pair with significant difference for the parameters FEV1/FVC and FEV1 (% of predicted) and two group pairs for the residual volume (% of predicted). The CT emphysema index yields two distinguishable group pairs. For the parameters signal homogeneity and areal texture retrieved from the reader study of the dark-field images, we obtain 3 group pairs which indicated significant variation. None of the evaluated parameter showed significant distinguishability for the group pair *absent–trace*. Table 3 and the box plots in figure 4B list and illustrate these findings.

**Table 3:** Study Evaluation. Emphysema assessment based on features observable in CT and dark-field images yield consistent findings. With respect to Fleischner emphysema grading, dark-field image features exhibit improved diagnostic performance in comparison to conventional clinical parameters.

| Overview of study results | Mean (95% confidence interval) |  |  |  | Minimum | Maximum |  |
| --- | --- | --- | --- | --- | --- | --- | --- |
| CAT™ | 10.0 (8.5, 11.4) |  |  |  | 0.0 | 30.0 |  |
| FEV1/FVC<sup>I</sup> | 0.77 (0.74, 0.79) |  |  |  | 0.38 | 0.99 |  |
| FEV1 % of predicted<sup>I</sup> | 96.3 (90.3, 102.2) |  |  |  | 23.5 | 147.8 |  |
| Residual volume % of predicted<sup>I</sup> | 120.1 (110.7, 129.6) |  |  |  | 55.2 | 231.9 |  |
| DLCO SB % of predicted (N=42) | 86.5 (79.7, 93.2) |  |  |  | 30.0 | 129.1 |  |
| CT emphysema index [%]<sup>II</sup> | 3.3 (1.4, 5.1) |  |  |  | 0.0 | 39.4 |  |
| Dark-field<sup>I</sup> -ln( V/ V<sub>0</sub>)<sup>III</sup> | 0.29 (0.27, 0.31) |  |  |  | 0.06 | 0.50 |  |
| Dark-field based reader ratings<sup>IV</sup> |  |  |  |  |  |  |  |
| Rated dark-field signal | 4.21 (4.02, 4.41) |  |  |  | 1.83 | 5.80 |  |
| Emphysema severity level | 0.98 (0.72, 1.25) |  |  |  | 0.00 | 4.00 |  |
| Homogeneity level | 3.61 (3.49, 3.73) |  |  |  | 1.93 | 4.00 |  |
| Texture level | 0.50 (0.36, 0.65) |  |  |  | 0.00 | 2.50 |  |
| Study results by visual emphysema grades<sup>V</sup> | Absent | Trace | Mild | Moderate | Confluent, Adv.destr. | H-Value | P-Value |
| Number of subjects | 35 | 19 | 9 | 7 | 7 |  |  |
| Clinical Parameters |  |  |  |  |  |  |  |
| CAT™ | 8.60<br>(±5.88)<br>8.00 | 8.53<br>(±6.59)<br>8.00 | 9.89<br>(±3.14)<br>11.00 | 12.43<br>(±5.80)<br>13.00 | 18.29<br>(±7.93)<br>20.00 | 11.98 | 0.0175 |
| FEV1/FVC<sup>I</sup> | 0.81<br>(±0.09)<br>0.81 | 0.81<br>(±0.07)<br>0.79 | 0.72<br>(±0.08)<br>0.71 | 0.67<br>(±0.11)<br>0.72 | 0.60<br>(±0.17)<br>0.55 | 25.63 | <0.0001 |
| FEV1 % of predicted<sup>I</sup> | 104.04<br>(±23.43)<br>105.90 | 105.93<br>(±17.83)<br>103.40 | 88.46<br>(±19.34)<br>83.10 | 79.87<br>(±27.47)<br>86.00 | 57.69<br>(±23.12)<br>54.30 | 21.94 | 0.0002 |
| Residual volume % of predicted<sup>I</sup> | 109.42<br>(±37.60)<br>106.60 | 105.49<br>(±22.20)<br>104.00 | 120.67<br>(±28.31)<br>117.00 | 155.67<br>(±44.36)<br>156.50 | 177.33<br>(±53.51)<br>208.80 | 16.20 | 0.0028 |
| CT emphysema index [%]<sup>II</sup> | 1.49<br>(±3.12)<br>0.13 | 0.93<br>(±2.54)<br>0.15 | 0.95<br>(±1.18)<br>0.43 | 2.23<br>(±4.00)<br>0.64 | 22.34<br>(±15.84)<br>29.94 | 21.63 | 0.0002 |
| Dark-field based reader evaluation <sup>IV</sup> |  |  |  |  |  |  |  |
| Emphysema severity level<br>no evidence (0), beginning- (1), mild- (2), moderate- (3), and severe (4) | 0.47<br>(±0.50)<br>0.40 | 0.45<br>(±0.62)<br>0.20 | 1.24<br>(±0.80)<br>1.40 | 2.11<br>(±1.30)<br>2.60 | 3.54<br>(±0.46)<br>3.60 | 31.86 | <0.0001 |
| Homogeneity level<br>very inhomogeneous (1), moderately inhom. (2), mildly inhom./mostly homogeneous (3), hom. (4) | 3.85<br>(±0.13)<br>3.90 | 3.86<br>(±0.11)<br>3.90 | 3.60<br>(±0.32)<br>3.73 | 2.93<br>(±0.67)<br>2.97 | 2.40<br>(±0.30)<br>2.40 | 36.26 | <0.0001 |
| Texture level<br>no texture (0), subtle stains (1, <5 mm), medium stains (2, 5-10 mm), and large stains (3, >10mm) | 0.21<br>(±0.21)<br>0.13 | 0.19<br>(±0.18)<br>0.13 | 0.60<br>(±0.50)<br>0.40 | 1.27<br>(±0.64)<br>1.10 | 1.90<br>(±0.44)<br>1.87 | 36.92 | <0.0001 |
| Pair-wise comparison<sup>VI</sup> | Absent-Trace |  | Trace-Mild | Mild-Moderate | Moderate-Confluent, Adv.destr. |  |  |
| CAT™ | 0.4638 |  | 0.1326 | 0.1428 | 0.1114 |  |  |
| FEV1/FVC<sup>I</sup> | 0.2096 |  | **0.0027** | 0.3355 | 0.2214 |  |  |
| FEV1 % of predicted<sup>I</sup> | 0.4639 |  | **0.0194** | 0.4162 | 0.0626 |  |  |
| Residual volume % of predicted<sup>I</sup> | 0.4711 |  | **0.0472** | **0.0452** | 0.2615 |  |  |
| CT emphysema index [%]<sup>II</sup> | 0.2514 |  | **0.0383** | 0.1449 | **0.0053** |  |  |
| Emphysema severity level | 0.2680 |  | **0.0048** | 0.0614 | **0.0051** |  |  |
| Homogeneity level | 0.3959 |  | **0.0089** | **0.0113** | **0.0482** |  |  |
| Texture level | 0.4101 |  | **0.0084** | **0.0245** | **0.0276** |  |  |
| <sup>I</sup> For patients with FEV1/FVC <0.7 measurement results post bronchodilator administration were utilized for the statistical analysis. |  |  |  |  |  |  |  |
| <sup>II</sup> Percentage of the segmented lung that exhibits density values below -950 HU. |  |  |  |  |  |  |  |
| <sup>III</sup> Value for one patient corresponds to the average signal value of all pixels within the lungs. |  |  |  |  |  |  |  |
| <sup>IV</sup> Emphysema severity rating was obtained by calculating the mean over all five readers. For the parameters rated dark-field intensity, homogeneity and texture level, the average over all six subregions and all readers was used for the evaluation. |  |  |  |  |  |  |  |
| <sup>V</sup> Values correspond to mean (standard deviation), median within respective visual emphysema grade. Critical H-value for a confidence level of 0.95 in Kruskal-Wallis test: H=9.49. |  |  |  |  |  |  |  |
| <sup>VI</sup> P-values from Mann-Whitney-U test. P-values <0.05 were considered to indicate significant difference (bold). |  |  |  |  |  |  |  |

Consistently, subjects graded with *absent* or *trace* of emphysema in the visual CT evaluation were generally rated with lower severity levels based on the dark-field images. The ratings steadily increase with *mild, moderate, confluent* and *advanced destructive* emphysema. Larger variations in diagnostic confidence with respect to this classification were not found (*absent*: 3·6, trace: 3·6, *mild*: 3·5, *moderate*: 3·7, *confluent* and *advanced destructive*: 3·9, all mean within Fleischner groups).

## Discussion

These results demonstrate clearly that dark-field chest radiography can – after 12 years of research since its invention on the optical bench – successfully be translated to human applications using readily available X-ray imaging hardware and acceptable exposure doses. Furthermore, the first patient results confirm that X-ray dark-field chest imaging is capable of detecting structural impairments associated with COPD, which remains challenging using only conventional chest X-rays. As the lung’s diffusion capacity strongly depends on their alveolar surface area, which in turn affects small angle scattering of X-rays, the dark-field signal encodes spatially resolved information about lung health. This assigns the dark-field signal a functional character in terms of chest imaging, which is confirmed by the good correlation with pulmonary function testing. However, these findings also indicate that spirometry cannot be associated entirely with the dark-field signal. A possible explanation might be the limited ability of pulmonary function testing to detect initial structural changes. Also, airflow limitation may express obstruction of the small airways without the presence of emphysema.^14^ In addition, several groups have demonstrated that many smokers without traditional spirometric obstruction experience symptoms and structural changes (emphysema) similar to those seen in spirometrically diagnosed COPD.^34,35^ In a study by Lynch et al., some degree of emphysema at CT was found in 562 (44%) of 1285 smokers with no spirometric abnormality.^36^ Furthermore, Oh et al. demonstrated recently that visual presence of emphysema in CT data from this population predicts progressive emphysema, lung function loss, and greater mortality.^37^ Hence, it is not surprising to observe a gradual decrease of the dark-field signal in few patients exhibiting a normal FEV1/FVC ratio.

In contrast to pulmonary function testing, dark-field radiography as an imaging technique allows for a regional evaluation of the lungs’ structural condition. This feature might be utilized for an improved treatment monitoring, phenotyping, or intervention planning.

Compared to the CT emphysema index, where pathological classification is binary, the dark-field signal allows for a gradual assessment, as it characterizes structural properties of lung parenchyma. This is reflected in the improved capability to register the diffusion capacity (DLCO SB) in our study. Apart from that, the thresholding approach used to obtain the emphysema index is very sensitive to noise in the reconstructed images (and thus also to the used reconstruction algorithm, slice thickness, and dose), since affected and healthy tissue exhibit similar density values.^38,39^ For this reason, the differentiation of early disease stages using the CT emphysema index is limited.^36^ This is also apparent from our findings for the tendency across the Fleischner grades where all groups until *moderate* emphysema yield mean index values below the critical value of 6%.

Visual features (signal homogeneity and texture) observable in the dark-field images appear to provide greater diagnostic value than conventional emphysema characterizing parameters when taking the Fleischner grading scheme as a reference standard.

In direct comparison, dark-field images and visual evaluation of CT images yield consistent findings regarding emphysema diagnosis. This is not surprising considering that the Fleischner grade assignment is based on the size and distribution of regional density reduction originating from parenchymal impairments. The associated changes in the alveolar microstructure also induce local signal variations in the dark-field image due to reduced small-angle scattering in affected regions. In turn, this affects the homogeneity and areal texture of the dark-field signal within the lung regions.

Drawing conclusions about the diagnostic performance of the dark-field images with respect to early disease stages based on this data is restricted as the reference standard itself might have limitations detecting initial changes. We find that the Fleischner scale does not deliver a significant correlation to the diffusion capacity if only emphysema grades from *absent* to *mild* are regarded, while the dark-field signal intensity still showed a moderate correlation for these groups in the examined collective. This might indicate an ambiguous interpretation of the true lung condition using CT at initial stages, limiting validation of diagnostic performance. This question has to be addressed in a longitudinal study with a larger cohort.

The main limitation of this study lies in the relatively small number of included patients. The presented findings have to be confirmed by a large cohort of smokers with or without airflow obstruction and a small group of healthy non-smokers who all underwent both chest CT and dark-field chest X-rays. In addition, an absence of dark-field signal decrease in patients with non-emphysematous obstructive lung disease, such as obliterative bronchiolitis and/or severe persistent asthma, should contribute to measure the specificity.

Emphysema-associated reduction of the respiratory surface has a negative impact on the lungs’ diffusion capacity and is generally irreversible. However, treatment options exist and can delay disease progression. Early disease detection is therefore crucial for an effective treatment and consequently an improved quality of life.^32^ At the same time, COPD and emphysema are highly unrecognized conditions.^34,35^ Due to the current absence of disease-modifying therapies, the role of COPD screening programs, which could resolve this underdiagnosis, is discussed controversially.^35^ On the other hand, it has been reported that relevant imaging data with a focus on early disease stages is required for therapeutic development.^15^

With effective dose values remarkably below those of low-dose CT (only ∼2%), the dark-field technique could contribute to an increased acceptance towards screening programs. Providing detailed information on lung micromorphology, dark-field chest radiography could serve as a low-dose imaging tool enabling periodic examinations to resolve the prevalent condition of underdiagnosed COPD. In the context of recently reported advances in regenerative therapeutics,^40^ dark-field imaging might support further translational research in this field and might at a later stage be integrated in disease management concepts for the monitoring of treatment response.

With this work, we report the first results of X-ray dark-field chest imaging applied in humans and demonstrate that this technology enables the detection and quantification of pulmonary emphysema. As dark-field chest imaging is not limited to COPD, further translational studies on other lung pathologies, such as fibrosis, pneumothorax, lung cancer, or pneumonia (including COVID-19) are of great interest as well.^8,9,11,12^ Based on the reported results, we believe that X-ray dark-field chest imaging can contribute substantially to improve the detection, diagnosis, and thus treatment and care of pulmonary disorders.

## Data Availability

All data required to evaluate the conclusions of our study are presented in the manuscript.

## Acknowledgments

We acknowledge gratefully Florian Gassert and Jannis Bodden for performing parts of the visual image analysis, Felix Meurer, Yannik Leonhardt, Christina Müller-Leisse, Martin Renz, Nadja Meissner, and Angelika Kammermeier for help with patient handling, Margarete Kattau, David Jany, Hanns-Ingo Maack, Hendrik van der Heijden, Jens von Berg, Klaus-Jürgen Engel, Bernd Lundt, Sven Prevrhal, Karsten Rindt, Roland Proksa, Michael Heider, Bernhard Haller, Pascal Meyer, and Jürgen Mohr for assistance during the hardware- and software-development for the demonstrator system, and Peter Noël for help with the approval process.

## Contributors

All authors elaborated the study design. K.W., W.N., F.D.M., M.F., T.U., R.S., A.G., B.G., T.K., A.Y., T.P., B.R., J.H., and F.P. developed hardware and control software of the demonstrator system. K.W., W.N., F.D.M., M.F., T.U., R.S., T.K., A.Y., T.P., J.H., and F.P. developed the data processing algorithms. A.F., G.Z., and D.P. interpreted the results from pulmonary function tests. A.F., A.S., D.P., M.M., E.R., and P.G planned the radiological interpretation of imaging data. A.F., A.S., and D.P. performed the visual image analysis. K.W. and A.F. performed the statistical analysis. J.H., M.M., E.R., and F.P. supervised the project. K.W. and A.F. wrote the original draft and revised the manuscript with input from all authors. All authors approved the manuscript.

## Data sharing

All data required to evaluate the conclusions of our study are presented in the manuscript.

## Supplementary material

The supplementary material covers further information on the CT measurements, statistical data analysis, and our demonstrator system including hardware, its clinical operation, and signal correction procedures.

We additionally present an overview about the study results of two additional participants with opposite smoking habits, one without pulmonary impairment and one with severe emphysematous destruction.

An exemplary video sequence of a patient scan is also available online.

## Supplementary material

The online material provides further information on our demonstrator system, CT evaluation, and statistical data analysis. Furthermore, we present the study results of two additional participants with opposite smoking habits, one without pulmonary impairment and one with severe emphysematous destruction.

### Chest X-ray dark-field demonstrator system

#### Hardware

The high-aspect-ratio grating structures were manufactured utilizing X-ray lithography and subsequent gold electroplating (microworks GmbH, Germany). Due to current limitations in grating fabrication, typical sizes currently achieved with this technique range in the order of some centimeters. To extend the coverage suitable to image an entire human thorax, a combination of grating tiling^41^ and interferometer scanning^26,27^ was established for the demonstrator system. As it is the grating most distant from the source, it is the analyzer grating G_2_ that limits the size of the active area usable for acquisition. It is assembled from six individual grating tiles (7·0×6·5 cm^2^) to entirely cover the field of view in horizontal direction. By moving the interferometer relative to the imaging system during acquisition, the desired coverage in the vertical direction is achieved. In addition, relative phase shifts between analyzer grating and the high-frequency intensity pattern generated by G_1_ were encoded in the vertical coordinate of the active area. By that, a sinusoidal data set carrying attenuation and dark-field information can be sampled within one pixel during the scan movement. The relative shifts arise in the form of horizontal moiré fringes which are depicted in figure S1 by an exemplary single exposure without patient. Over the entire field of view, we obtain a mean visibility of about 31%.

**Figure S1:**
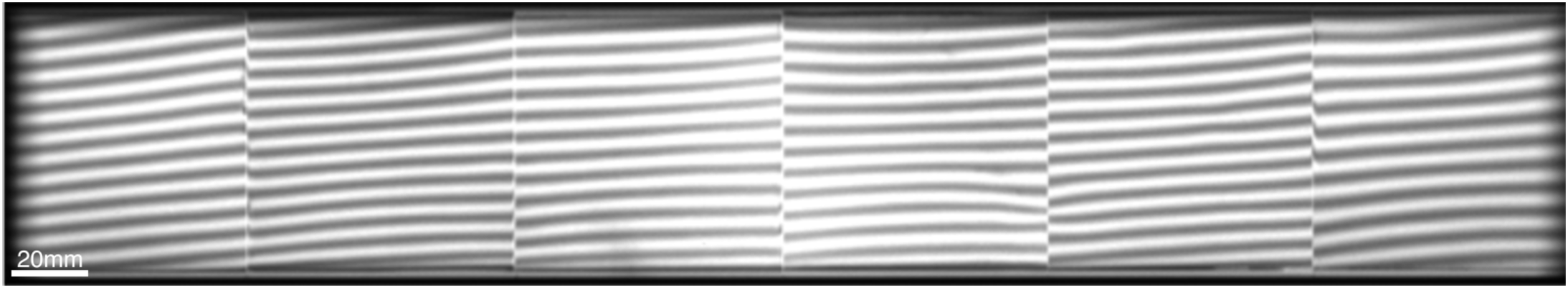
Single raw exposure without patient. The G_2_ is stitched together from six individual high aspect ratio gold grating tiles forming the active area of about 42×6·5 cm^2^. With 0·5 mm, the gap size between the tiles slightly exceeds the image pixel size. By scanning the active area in vertical direction, a field of view of about 37×37 cm^2^ with respect to the patient plane is achieved. The moiré fringes serve as sampling pattern for the attenuation and the dark-field signal.

In order to avoid shadowing induced by the binary grating structures, all gratings are adapted to the divergent beam shape by cylindrical bending with respect to the corresponding distance to the focal spot of the X-ray source. In addition, the scan movement is performed following an arc, also centered on the focal spot, to preserve the incident beam angle onto the gratings throughout the scan. The grating arrangement is mounted on an aluminum frame construction that is attached to a spindle drive performing the scan movement. The connection to the drive is realized as a sliding hinge to provide the required degrees of freedom for the rotational scan movement.

After a short acceleration phase, as soon as the active area reaches the field of view of the imaging system, the source (MRC 200 0508, Royal Philips, The Netherlands) is triggered to emit X-ray pulses with a repetition rate of 30 Hz. Simultaneously, the flat-panel detector (Pixium FE 4343, Trixell, France) acquires images with an exposure time of about 17 ms per frame. If utilizing the maximal achievable field of view, a total of 195 frames are acquired. A video sequence (video 1) playing back raw detector images of an actual patient scan in real time is available online.

#### Clinical operation

The operation of the scanner is comparable to the one of a conventional medical radiography system. Prior to the scan, the medical-technical radiology assistant explains the procedure to the patient and adjusts the field of view as well as the patient’s position. After moving the interferometer to the starting position and leaving the examination room, the operator can trigger the acquisition via a control panel. For the duration of the scan, the patient remains positioned in the optical path of the scanner, holding breath. To properly synchronize breath-hold and acquisition, the patient obtains automatically-triggered audio commands during the acceleration phase of the interferometer and after the acquisition. In order to reduce displacement artifacts, the scan is performed in upwards direction, as premature breathing (if prevalent) is expected to be more likely to occur towards the end of the acquisition. At that time, the active grating area has already passed the diaphragm region where larger shifts of tissue are expected.

#### Corrections and signal calibrations

In addition to an air scan normalization, corrections on the dark-field and attenuation images are applied in order to account for Compton scattering and beam hardening.

Secondary photons that originate from Compton scatter can be considered a diffuse illumination that increases the mean value of the fringe pattern, but not its amplitude. This results in a decreased visibility and thus has a negative influence on the measurement of the dark-field signal. Although the high-aspect-ratio G_2_ grating already removes a majority of these photons, a significant part, primarily scattered in a direction parallel to the grating lamellae, still reaches the detector. To account for this effect, a commercially available software (SkyFlow, Royal Philips, The Netherlands) for the estimation of Compton scattering, based on Monte Carlo simulations and a preliminary attenuation image, was adapted to the dark-field demonstrator system and is utilized to correct the raw images before data processing.

The beam hardening correction is applied to the dark-field images to account for spectral differences between patient and reference scan^42^. Hereby, calibration data obtained from a series of visibility measurements with a varying number of tissue-equivalent absorbing sheets introduced into the beam path is used to correct the air-scan visibility. The attenuation image is used to generate a correction map, based on the assumption of similar modification of the X-ray spectrum in the calibration and the patient scan.

Image post-processing including histogram adaptation and edge enhancement using a commercially available software (UNIQUE, Royal Philips, The Netherlands) is performed on the attenuation image to obtain an image quality comparable to conventional systems. The data given in Table 2 was extracted prior to this post-processing. In the dark-field image, pixel lines coinciding with tiling gaps (∼0·5 mm) are replaced by interpolation between neighboring pixels. This step can be omitted for the attenuation signal as the gaps are corrected by the air scan normalization.

Determining the illumination during the scan, the collimated active area moves across one pixel in roughly 0·8 sec. Tissue displacement during this time interval, in particular occurring in the heart and diaphragm region, can induce artefacts in the reconstructed images. To reduce these artefacts, a correction algorithm is applied that is based on virtually curtailing the sampling of affected regions.

In order to obtain uniform noise variance independent from the sample’s transmission, adaptive denoising is applied to the dark-field image.

### Quantitative CT evaluation

Figure S2 shows screenshots of the emphysema evaluation software (IntelliSpace, Royal Philips, The Netherlands) for the patient depicted in figure 2 of the main manuscript. The histogram illustrates the density distribution within the lung region. All voxels featuring density values below the threshold are assigned to be emphysematous, 11·5% in this case. The upper-lobe impairment predominance revealed in the dark-field image (figure 2B) is also indicated by the sectional evaluation by lung lobe: Right upper lobe (RUL): 21·1%; right middle lobe (RML): 13·7%; right lower lobe (RLL): 3·5%; left upper lobe (LUL): 14·3%; left lower lobe (LLL): 3·4%.

**Figure S2:**
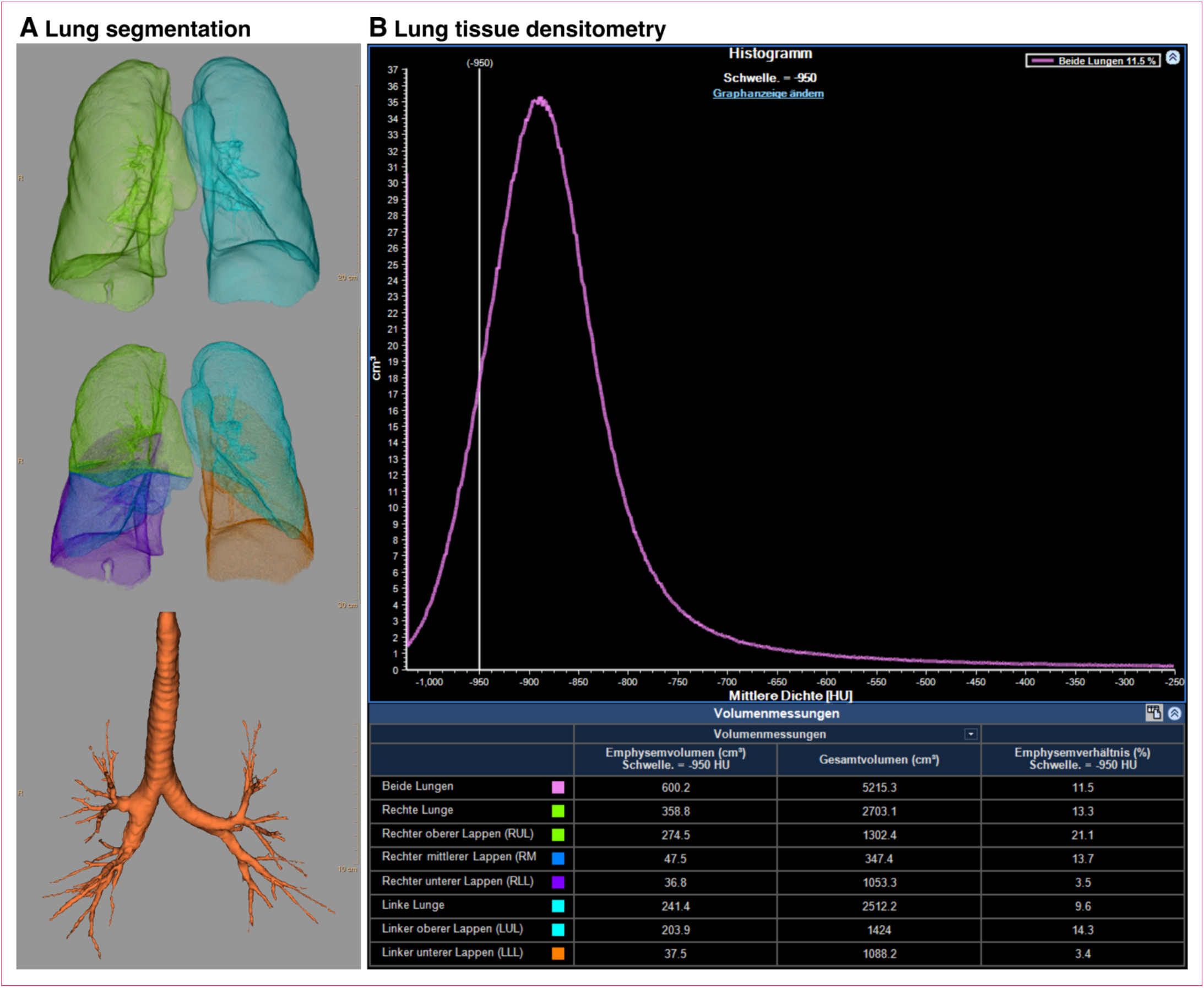
Screenshots of CT emphysema evaluation for the patient presented in figure 2. (A) shows the result of the segmentation of lungs, lobes, and bronchi. In (B), the density distribution within the segmented lung and a detailed listing of the emphysema index (rightmost table column), subdivided into lung side and lobes are depicted. The vertical line in the histogram denotes the threshold (−950 HU) for the evaluation. In the present case, 11·5% of the entire lung volume is classified as emphysematous.

### Statistical analysis

To compare the dark-field signal strength with the reader ratings, an individual mask for every patient was generated by manually following the boundary of the lungs in the attenuation image. Similar to the reader study, left and right lung were split in three parts of (approximately) equal height between apex and costodiaphragmatic recess. As both signal channels originate from the same data set, the pixel information is perfectly registered, and the mask coordinates can be transferred directly to the dark-field channel. As an example, figure S3 shows the mask for the patient in figure 1B–D of the main manuscript. For the mask generation, calculations of the correlation coefficients, and data handling, open-source software frameworks (NumPy, PIL, matplotlib, SciPy, Pandas, Python) were utilized.

**Figure S3:**
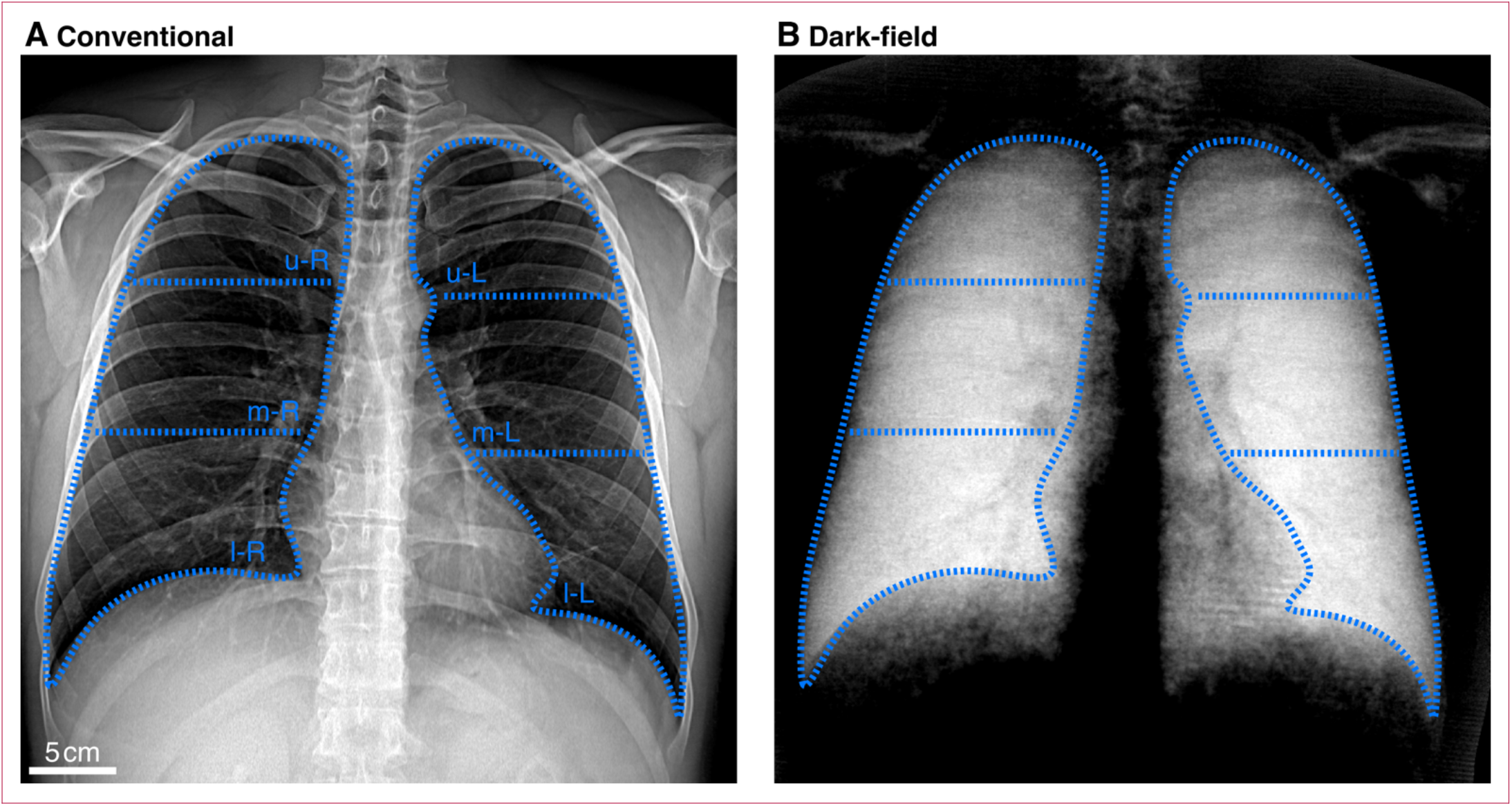
Exemplary lung masks generated manually for the statistical evaluation. Conventional (A) and dark-field (B) images of the patient depicted in Figure 1 with superimposed lung masks. The masks were generated by manually tracing the boundary of the lungs in the attenuation image and splitting each lung into an upper, middle, and lower part. Heart and diaphragm regions were omitted. In order to obtain a reference measure for comparison with the reader ratings, the average signal from all coordinates lying within the boundary was calculated.

### Additional cases

In the following, we present two additional cases from our study. Figure S4 provides an overview of all study data for a patient (male, 71 years, BMI 29·2) without smoking history. The attenuation-based radiograph (figure S4A) provides no conspicuous findings with regard to pulmonary emphysema, and the air-flow curve as well as the spirometric parameters (figure S4C) indicate normal pulmonary function. No secondary measurement with prior bronchodilator administration was performed, as no signs suggesting pulmonary obstructions were prevalent. The dark-field image (figure S4B) exhibits a homogeneous signal over the entire lung region. With a mean value of 0·28, however, the signal average is slightly reduced compared to those obtained for younger, healthy patients. This slight reduction also appears to be implied by the findings in CT densitometry (figure S4D) indicating a slightly elevated volume fraction (3·9%), with lower density values distributed homogeneously over the entire lung.

**Figure S4:**
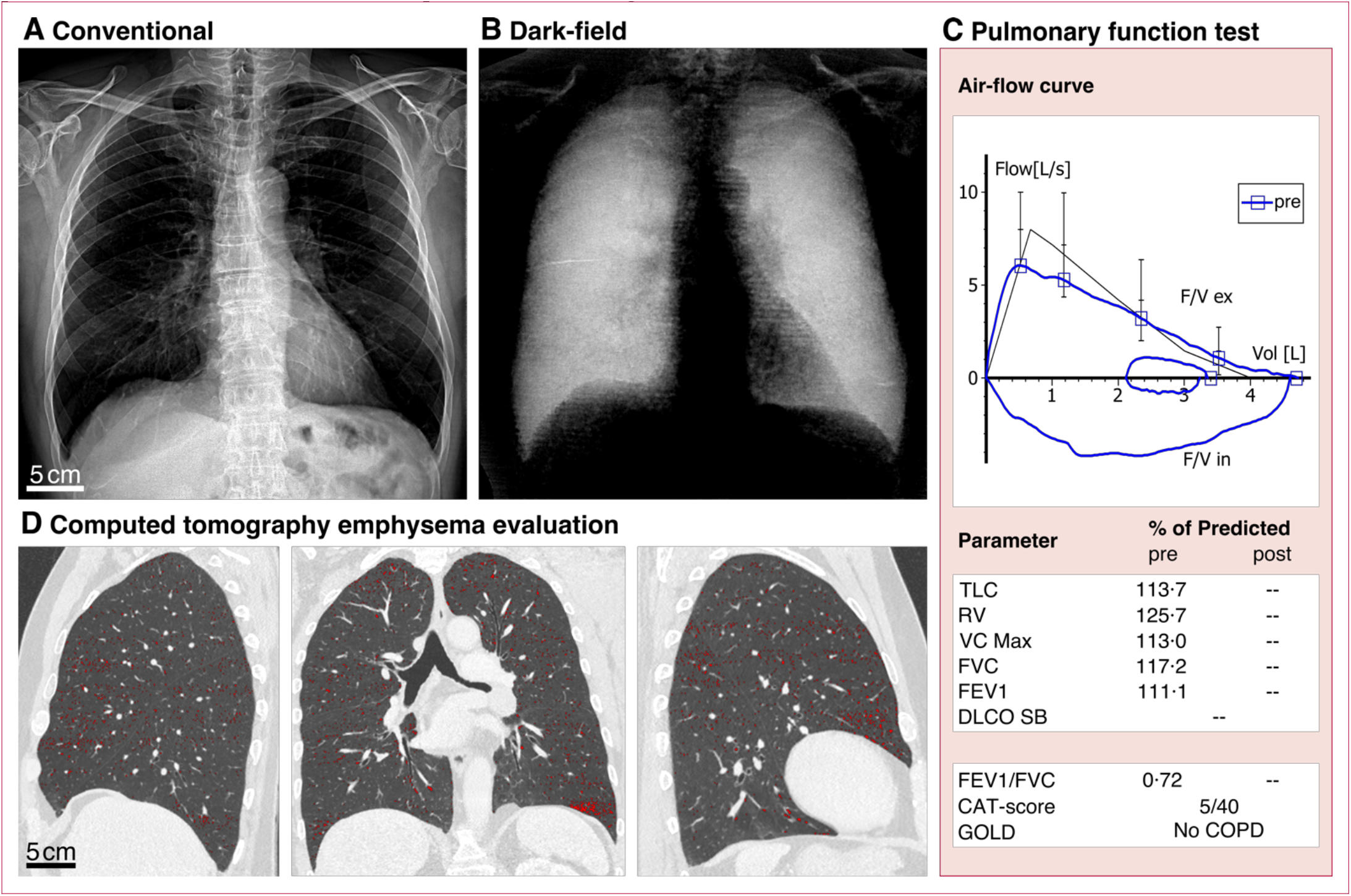
Clinical study results of a patient without COPD. Conventional attenuation (A) and dark-field (B) images of a patient (male, 71 years, BMI 29·2, non-smoker) without clinical indications of COPD. The dark-field image shows a homogeneous signal distribution over the lung with a slightly decreased mean value of 0·28, which might be an indication for a beginning impairment of the respiratory surface. For the attenuation signal, an average value over the entire lung of 2·74 was measured. With an inconspicuous air-flow curve and normal spirometric parameters (C), a healthy condition of the lung is indicated. CT densitometry yields an emphysema ratio of 3·9%, with detected areas being finely distributed over the entire lung, as depicted in the CT slices (D) where corresponding pixels are marked in red.

The second patient (male, 65 years, BMI 21·0) is a heavy smoker with a history of 80 pack-years, suffering from severe pulmonary emphysema. In figure S5, the respective study results are depicted. The attenuation image (figure S5A) shows enlarged intercostal spaces, a long barrel thorax shape including an elongated heart region, and a flat diaphragm, which all are advanced-stage, secondary symptoms of an emphysema. No significant contrast variation is observable. Indicating a substantial reduction of respiratory surface, low dark-field signal values (figure S5B) are obtained over the entire lung region. Towards apical regions appearing with inhomogeneous signal texture, a stronger manifestation of emphysematous destruction is revealed. Classifying the patient in GOLD-group 4, spirometry (figure S5C) yields a highly obstructed pulmonary condition that is preserved after bronchodilation. Substantial impairment is also reflected by a high volume-fraction (26%) featuring reduced density values in the CT reconstruction. Similar to the dark-field radiographs, a stronger emphysema manifestation can be identified in upper lung regions in the CT images (figure S5D). The regional prevalence of reduced density in the CT images is in good accordance with the signal distribution in the dark-field image.

**Figure S5:**
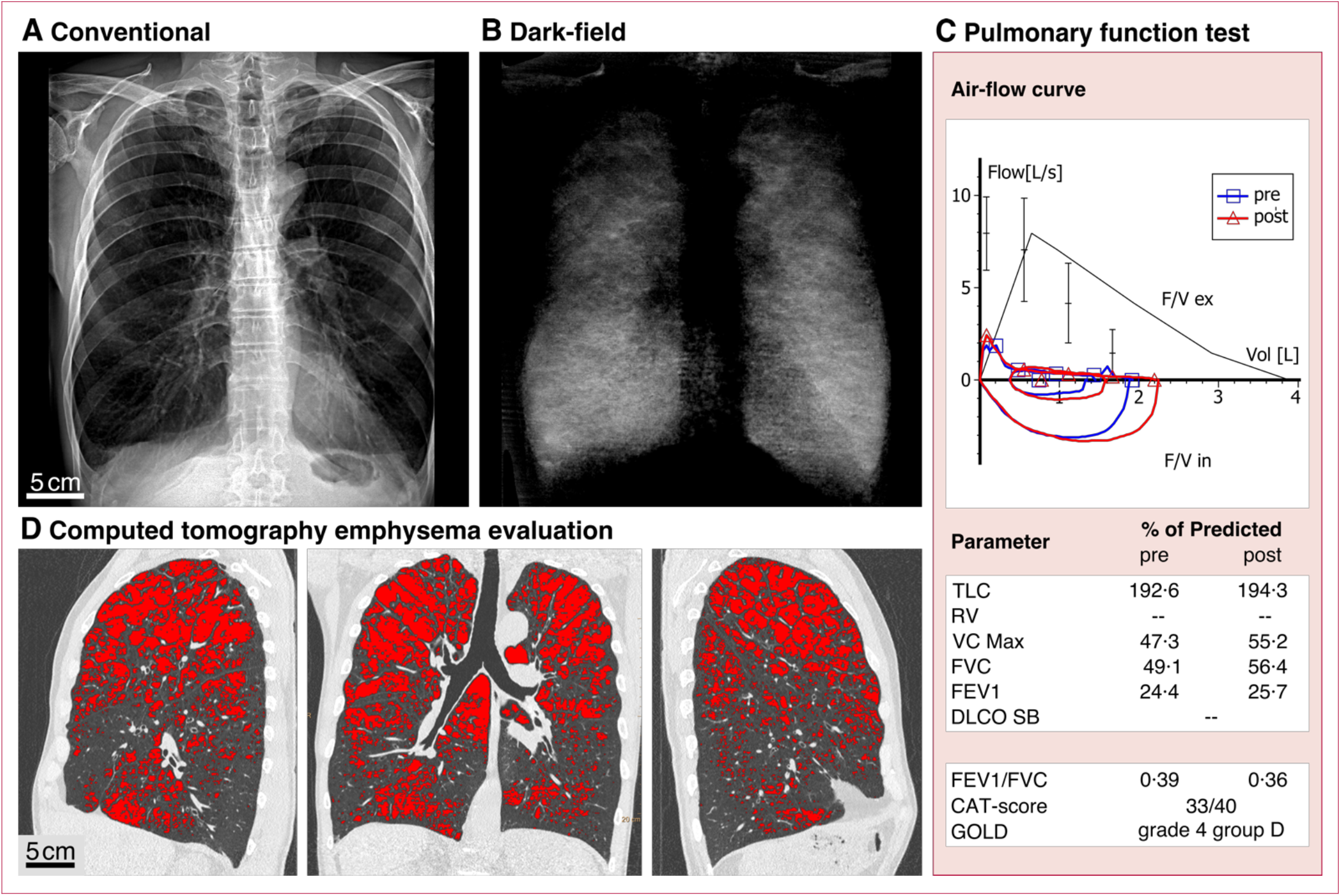
Study results of a patient graded with GOLD stage 4. Conventional attenuation (A) and dark-field (B) images of a patient (male, 65 years, BMI 21·0, 80 pack-years) suffering from severe emphysema affecting the entire lung, with stronger manifestation in upper regions. Reduced dark-field values indicate a degraded alveolar network with a decreased number of air-tissue interfaces. Over the entire pulmonary region, a mean value of 0·07 was measured for the dark-field and 2·31 for the attenuation. Spirometry (C) yields an obstructive condition of the lung with strongly deviating parameters that are preserved after administration of a bronchodilator. CT densitometry yields an emphysema index of 26%. The corresponding voxels in the CT slices (D) are marked in red. Dark-field and CT images yield consistent information with regard to parenchymal condition. Note that, this patient was not included in the statistical evaluation, as he underwent a native CT examination.

Caption for video 1

**Video 1: Exemplary video sequence of a patient scan.** The sequence comprises raw images of an actual patient scan and was generated from the raw pixel data as obtained from the flat-panel detector. The acquisition frequency (30 Hz) was set as frame rate of the movie, playing back the acquisition in real time. In order to retrieve the conventional attenuation and dark-field images, the moiré pattern overlaying the patient’s thorax is mapped onto a sinusoidal model in subsequent processing steps. At the contour of the heart, slight displacements induced by the heartbeat can be recognized. The scan movie shows the acquisition of the patient presented in figure 1B-D of the main manuscript.

## References

1. Pfeiffer F, Bech M, Bunk O, et al. Hard-X-ray dark-field imaging using a grating interferometer. Nat Mater 2008; 7: 134–37.

2. Schleede S, Meinel FG, Bech M, et al. Emphysema diagnosis using X-ray dark-field imaging. Proc Natl Acad Sci 2012; 109: 17880–85.

3. Yaroshenko A, Meinel FG, Bech M, et al. Pulmonary emphysema diagnosis with a preclinical small-animal X-ray dark-field scatter-contrast scanner. Radiology 2013; 269: 427–33.

4. Bech M, Tapfer A, Velroyen A, et al. In-vivo dark-field and phasecontrast X-ray imaging. Sci Rep 2013; 3: 3209.

5. Bech M, Schleede S, Potdevin G, et al. Experimental validation of image contrast correlation between ultra-small-angle X-ray scattering and grating-based dark-field imaging using a laser-driven compact X-ray source. Photon Laser Med 2011; 1: 47–50.

6. Meinel FG, Yaroshenko A, Hellbach K, et al. Improved diagnosis of pulmonary emphysema using in vivo dark-field radiography. Invest Radiol 2014; 49: 653–58.

7. Hellbach K, Yaroshenko A, Meinel FG, et al. In vivo dark-field radiography for early diagnosis and staging of pulmonary emphysema. Invest Radiol 2015; 50: 430–35.

8. Yaroshenko A, Hellbach K, Yildirim AÖ, et al. Improved in vivo assessment of pulmonary fibrosis in mice using X-ray dark-field radiography. Sci Rep 2015; 5: 17492.

9. Hellbach K, Yaroshenko A, Willer K, et al. Facilitated diagnosis of pneumothoraces in newborn mice using X-ray dark-field radiography. Invest Radiol 2016; 51: 597–601.

10. Yaroshenko A, Pritzke T, Koschlig M, et al. Visualization of neonatal lung injury associated with mechanical ventilation using X-ray dark-field radiography. Sci Rep 2016; 6: 24269.

11. Scherer K, Yaroshenko A, Bölükbas DA, et al. X-ray dark-field radiography – in-vivo diagnosis of lung cancer in mice. Sci Rep 2017; 7: 402.

12. Hellbach K, Meinel FG, Conlon TM, et al. X-ray dark-field imaging to depict acute lung inflammation in mice. Sci Rep 2018; 8: 2096.

13. World health organization (WHO). The top 10 causes of death. 2018. https://www.who.int/news-room/fact-sheets/detail/the-top-10-causes-of-death (accessed 2020 April 21).

14. Labaki WW, Martinez CH, Galbàn CG, et al. The role of chest CT in the evaluation and management of patients with chronic obstructive pulmonary disease. Am J Respir Crit Care Med 2017; 196: 1372–79.

15. Han MK. Should chest CT be part of routine clinical care for COPD? no. CHEST 2018; 154: 1278–81.

16. Mettler FA, Huda W, Yoshizumi TT, Mahesh M. Effective doses in radiology and diagnostic nuclear medicine: a catalog. Radiology 2008; 248: 254–63.

17. Larke FJ, Kruger RL, Cagnon CH, et al. Estimated radiation dose associated with low-dose chest CT of average-size participants in the national lung screening trial. Am J Roentgenol 2011;197: 1165–69.

18. Pratt PC. Role of conventional chest radiography in diagnosis and exclusion of emphysema. Am J Med 1987; 82: 998–1006.

19. Pfeiffer F, Weitkamp T, Bunk O, David C. Phase retrieval and differential phase-contrast imaging with low-brilliance X-ray sources. Nat Phys 2006; 2: 258–61.

20. Gromann LB, De Marco F, Willer K, et al. In-vivo X-ray dark-field chest radiography of a pig. Sci Rep 2017; 7: 4807.

21. Hellbach K, Baehr A, De Marco F, et al. Depiction of pneumothoraces in a large animal model using X-ray dark-field radiography. Sci Rep 2018; 8: 2602.

22. Willer K, Fingerle AA, Gromann LB, et al. X-ray dark-field imaging of the human lung – a feasibility study on a deceased body. PLoS One 2018; 13: e0204565.

23. De Marco F, Willer K, Gromann LB, et al. Contrast-to-noise ratios and thickness-normalized, ventilation-dependent signal levels in dark-field and conventional in vivo thorax radiographs of two pigs. PLoS One 2019; 14: e0217858.

24. Sauter AP, Andrejewski J, De Marco F, et al. Optimization of tube voltage in X-ray dark-field chest radiography. Sci Rep 2019;9: 8699.

25. Fingerle AA, De Marco F, Andrejewski J, et al. Imaging features in post-mortem X-ray dark-field chest radiographs and correlation with conventional X-ray and CT. Eur Radiol Exp 2019; 3: 25.

26. Kottler C, Pfeiffer F, Bunk O, Grünzweig C, David C. Grating interferometer based scanning setup for hard X-ray phase contrast imaging. Rev Sci Instrum 2007; 78: 043710.

27. Koehler T, Daerr H, Martens G, et al. Slit-scanning differential X-ray phase-contrast mammography: proof-of-concept experimental studies. Med Phys 2015; 42: 1959–65.

28. Donath T, Chabior M, Pfeiffer F, et al. Inverse geometry for grating-based X-ray phase-contrast imaging. J Appl Phys 2009; 106: 054703.

29. Coates AL, Peslin R, Rodenstein D, Stocks J. Measurement of lung volumes by plethysmography. Eur Respir J 1997;10: 1415–27.

30. Wanger J, Clausen JL, Coates A, et al. Standardization of the measurement of lung volumes. Eur Respir J 2005; 26: 511–22.

31. Lynch DA, Austin JHM, Hogg JC, et al. CT-definable subtypes of chronic obstructive pulmonary disease: a statement of the Fleischner Society. Radiology 2015; 277: 192–205.

32. Global Initiative for Chronic Obstructive Lung Disease (GOLD). Global strategy for the diagnosis, management, and prevention of chronic obstructive pulmonary disease 2019 report. 2019.

33. Johns DP, Walters JAE, Walters EH. Diagnosis and early detection of COPD using spirometry. J Thorac Dis 2014; 6: 1557–69.

34. Regan EA, Lynch DA, Curran-Everett D, et al. Clinical and radiologic disease in smokers with normal spirometry. JAMA Intern Med 2015; 175: 1539–49.

35. Martinez CH, Mannino DM, Jaimes FA, et al. Undiagnosed obstructive lung disease in the United States – associated factors and long-term mortality. Ann Am Thorac Soc 2015; 12:1788–95.

36. Lynch DA, Moore CM, Wilson C, et al. CT-based visual classification of emphysema: association with mortality in the COPDGene study. Radiology 2018; 288:859–66.

37. Oh AS, Strand M, Pratte K, et al. Visual emphysema at chest CT in GOLD stage 0 cigarette smokers predicts disease progression: results from the COPDGenestudy. Radiology 2020; 296:641–49.

38. Gierada DS, Bierhals AJ, Choong CK, et al. Effects of CT section thickness and reconstruction kernel on emphysema quantification relationship to the magnitude of the CT emphysema index. Acad Radiol 2010; 17: 146–56.

39. den Harder AM, de Boer E, Lagerweij SJ, et al. Emphysema quantification using chest CT: influence of radiation dose reduction and reconstruction technique. Eur Radiol Exp 2018; 2: 30.

40. Conlon TM, John-Schuster G, Heide D, et al. Inhibition of LTβR signalling activates WNT-induced regeneration in lung. Nature 2020; 588: 151–56.

## References (continued)

41. Schröter TJ, Koch FJ, Meyer P, et al. Large field-of-view tiled grating structures for X-ray phase-contrast imaging. Rev Sci Instrum 2017; 88: 015104.

42. Chabior M, Donath T, David C, et al. Beam hardening effects in grating-based X-ray phase-contrast imaging. Med Phys 2011; 38: 1189–95.

